# A systematic review on Multisystem Inflammatory Syndrome in Children (MIS-C) with COVID-19: Development of a scoring system for clinical diagnosis

**DOI:** 10.1101/2021.04.23.21255879

**Authors:** Suchitra V Surve, Shaini Joseph, Rahul K Gajbhiye, Smita D Mahale, Deepak N Modi

**Affiliations:** Department of Clinical Research, ICMR- National Institute for Research in Reproductive Health, J M Street, Parel, Mumbai 400012, INDIA; Department of Genetics, ICMR- National Institute for Research in Reproductive Health, J M Street, Parel, Mumbai 400012, INDIA; ICMR Emeritus Scientist, ICMR- National Institute for Research in Reproductive Health, J M Street, Parel, Mumbai 400012, INDIA; Department of Molecular and Cellular Biology Laboratory, ICMR- National Institute for Research in Reproductive Health, J M Street, Parel, Mumbai 400012, INDIA

**Keywords:** Children, COVID-19, Hyperinflammatory syndrome, Kawasaki disease, Multisystem involvement, SARS-COV-2

## Abstract

**Background:** There is growing evidence of Multisystem Inflammatory Syndrome in Children (MIS-C) resembling Kawasaki disease in children infected with SARS-CoV-2. The review was undertaken to evaluate the case definition, the spectrum of clinical presentations and current management practices in children with COVID-19 presenting with or without MIS-C.

**Methods:** The individual patient data from 119 studies accounting for 333 children were analyzed. We devised a scoring system as per WHO criteria to classify the patients as MIS-C or without MIS-C. A score of 3 was given for the presence of fever (>24h) and a score of 1 for lab-confirmed diagnosis of SARS-CoV-2. Additionally, a score of 1 was given for a) rash or conjunctivitis or muco-cutaneous inflammation signs, b) hypotension or shock, c) diarrhea, vomiting or abdominal pain, d) features of myocardial dysfunction as determined by abnormal eco-cardiography or elevated Troponin or N-terminal pro B-type Natriuretic Peptide (NT-proBNP), e) evidence of coagulopathy as evidenced by elevated levels of prothrombin time PT, partial thromboplastin time PTT or D-dimer, f) laboratory evidence of inflammation as determined by elevated erythrocyte sedimentation rate (ESR) or C-reactive protein (CRP) or procalcitonin. A negative score of (−3) was given when there was a diagnosis of sepsis, staphylococcal or streptococcal shock syndrome. Based on these criteria, a minimum score of 6 was essential to classify the child as MIS-C.

**Results:** Based on this score, 18% (52/289) of cases were identified to be MIS-C. A greater proportion of children with MIS-C had cardiac involvement (MIS-C 80% vs Non-MIS-C 20%) and gastrointestinal involvement (MIS-C 71% vs Non-MIS-C 12%). Lymphopenia was commonly reported in MIS-C (MIS-C 54.2% vs Non-MIS-C 29.7%). In addition to routine inflammatory markers, significantly greater proportion of children with MIS-C had elevated Ferritin, LDH, Fibrinogen and IL-6. Children with MIS-C were less likely to have respiratory symptoms like cough (MIS-C 25% vs Non-MIS-C 75%) and rhinorrhea (MIS-C 4% vs Non-MIS-C 22.8%). A greater proportion of children with MIS-C required intensive care and aggressive treatment; and mortality rates were also higher in MIS-C group (MIS-C 10% vs Non-MIS-C 1%).

**Conclusion:** The children with COVID-19 having cardiac and/or gastrointestinal involvement are more likely to develop MIS-C. The children with MIS-C have higher mortality rates. The scoring system developed herein will aid clinicians in patient diagnosis and timely management.

## Introduction

Children account for 1-5% of diagnosed novel coronavirus disease of 2019 (COVID-19) caused by severe acute respiratory syndrome coronavirus (SARS-CoV-2)^1–3^. Despite the initial assumption that children seem to be less severely affected by COVID-19^3^, there is growing evidence of diverse and unusual presentations of SARS-CoV-2 infections in the pediatric population.This is posing a huge challenge in the clinical management of children with COVID-19^4,5^.

Kawasaki disease (KD) is an acute febrile systemic vasculitis of unknown etiology occurring predominantly under the age of 5 years with a probable link with viral or bacterial illness^6–10^. It is hypothesized that an unknown trigger (most likely an infectious agent) leads to an exaggerated inflammatory response manifesting as KD. Previously, coronavirus infection has been reported to be associated with KD^10^. Case reports from Europe, USA and India have shown hyperinflammatory responses in pediatric COVID-19 which have overlapping features with KD^11–15^. However, whether these manifestations should be considered as a clinical spectrum of COVID-19 or as a result of a secondary infection triggered by SARS-CoV-2 is debatable. Nevertheless, for provisional reporting and surveillance, World Health Organization (WHO) and Center for Disease Control and Prevention (CDC) has categorized this condition as Multisystem Inflammatory Syndrome (MIS-C) related to COVID-19 in children and adolescents^16,17^.

The case definition proposed by WHO and CDC have notable differences. CDC proposes MIS-C as an individual aged <21 years presenting with fever ≥38.0°C for ≥24 hours, laboratory evidence of inflammation and evidence of clinically severe illness requiring hospitalization with multisystem (≥2) organ involvement (cardiac, renal, respiratory, hematologic, gastrointestinal, dermatologic or neurological); AND no alternative plausible diagnoses; AND positive for current or recent SARS-CoV-2 infection by RT-PCR, serology or antigen test; or COVID-19 exposure within the 4 weeks prior to the onset of symptoms. However, WHO case definition differs in terms of age group (includes children and adolescents <19 years), duration of fever (> 3days) and two of either rash or bilateral non-purulent conjunctivitis or muco-cutaneous inflammation signs (oral, hands or feet), hypotension or shock, myocardial dysfunction, pericarditis, valvulitis or coronary abnormalities, evidence of coagulopathy or acute gastrointestinal problems along with elevated markers of inflammation without any evidence of sepsis in presence of COVID-19 (RT-PCR, antigen test or serology positive) or contact with patients with COVID-19. Though the WHO criteria appear to be more specific concerning children, it needs refinement with regards duration of fever, the inclusion of other neurological and renal system involvement. Thus, there is a need to revisit the case definition and classification of MIS-C. Also, the clinical characteristics, the presenting symptoms and the outcome of children with MIS-C and the non-MIS-C needs to be determined.

With increasing numbers of case reports of MIS-C with SARS-COV-2, the available information is being collated systematically to create substantial learning in diagnosing and managing pediatric COVID cases^18,19^. These studies have highlighted that MIS-C is an integral component of pediatric infections with SARS-CoV-2. However, it is increasingly getting evident that the presentation of MIS-C in children appears diverse and there are differences in case definitions across various studies. This makes it harder for pediatricians to accurately define MIS-C patients in their clinical setup and epidemiologists to carry out detailed systematic reviews.

To address this, we carried out a systematic review that aimed at evaluating the case definition, the spectrum of clinical presentations and current management practices in children with COVID-19 presenting with and without MIS-C. The ultimate goal is to evolve a scoring system that would aid clinicians to classify MIS-C in their routine practice and identify the clinical characteristics of MIS-C.

## Methods

A systematic review (PROSPERO ID CRD42020185491) was carried out as per Preferred Reporting Items for Systematic reviews and Meta-analysis (PRISMA) guidelines.

### Search Strategy

The PubMed and Google Scholar literature database, journal databases, preprints servers and specialized resources like COVID-19 Research Database, LitCovid, CNKI database comprising of over 3900 Chinese journals were searched systematically using COVID-19 search terms: “coronavirus”, “COVID19”, “2019-nCoV”, “SARS-CoV-2”, “COVID-19”, “COVID” together with keywords commonly used to refer to pediatric population: “pediatric”, “neonate”, “newborn”, “new-born”, “infant”, “child*”, “children”. An additional search was also performed to check for articles on the multisystem inflammatory syndrome in children. The snowballing method was also applied to search for missed articles.

### Study selection and strategy for data extraction and synthesis

A pre-requisite for precise case definition requires sets of individual data. Since it is possible that early studies on children with COVID-19 may have reported the data but would have missed defining the case as MIS-C or conversely some studies may have over-interpreted the presentations as MIS-C, we chose to redefine the cases using a uniform criterion. Hence, we only included those studies where individual patient data were available as it allowed us to re-classify the patients uniformly.

All articles published between January 1, 2020 and June 1, 2020 were retrieved and filtered to remove duplicate entries. Studies published in other languages were included if translated by google translator. The articles of interest were selected in three steps, firstly based on article title, then article abstract and in the third step the full-text article was assessed. Original studies reporting primary individual patient data younger than 18 years of age and diagnosed with SARS-CoV-2 by laboratory method (PCR or antibody testing) were included. Studies reporting clinical guidelines, consensus documents, clinical trials, reviews, systematic reviews, clinical diagnoses of COVID-19 without laboratory confirmation were excluded. Primary data was extracted, cross-checked and any discrepancies were resolved.

### Development of the scoring system

To uniformly classify the patients as Multisystem Inflammatory Syndrome in Children (MIS-C) a scoring system was devised (Table 1). Herein a score of 3 was given for the presence of fever (>24h) and a score of 1 for a lab-confirmed diagnosis of SARS-CoV-2. Additionally, a score of 1 was given for a) rash or conjunctivitis or muco-cutaneous inflammation signs, b) hypotension or shock, c) diarrhea, vomiting or abdominal pain, d) features of myocardial dysfunction as determined by abnormal eco-cardiography or by elevated Troponin or N-terminal pro B-type Natriuretic Peptide (NT-proBNP) levels, e) evidence of coagulopathy as evidenced by elevated levels of prothrombin time PT, partial thromboplastin time PTT or D-dimer, f) laboratory evidence of inflammation as determined by elevated erythrocyte sedimentation rate (ESR) or C-reactive protein (CRP) or procalcitonin. A negative score of (- 3) was given when there was a diagnosis of sepsis, staphylococcal or streptococcal shock syndrome. Based on these criteria, a minimum score of 6 was essential to classify the child as MIS-C (Table 1).

**Table 1:** **Scoring and proportion of patients with and without various features of Multisystem Inflammatory Syndrome in Children (MIS-C). The p-value (by Chi-square test) compares MIS-C and Non-MIS-C groups**.

| Presentation | Score | MIS-C<br>n=52 (%) | Non-MIS-C<br>n=237 (%) | p-value |
| --- | --- | --- | --- | --- |
| Fever > 3 days | 3 | 52 (100) | 153 (64.6) | - |
| Lab diagnosis of SARS-CoV-2 | 1 | 52 (100) | 237(100) | - |
| Rash or conjunctivitis or mucocutaneous inflammation signs | 1 | 18 (34.6) | 10 (4.22) | <0.0001 |
| Hypotension or shock. | 1 | 39 (75) | 5 (2.11) | <0.0001 |
| Diarrhoea, vomiting, or abdominal pain | 1 | 37 (71.2) | 28 (11.9) | <0.0001 |
| Features of myocardial dysfunction, (abnormal eco-cardiography or elevated Troponin or N-terminal pro B-type Natriuretic Peptide (NT-proBNP) | 1 | 42 (80.8) | 3 (1.3) | <0.0001 |
| Evidence of coagulopathy (Elevated levels of prothrombin time PT, partial thromboplastin time PTT or D-dimer) | 1 | 29 (55.8) | 11 (4.7) | <0.0001 |
| Evidence of Inflammation Increased erythrocyte sedimentation rate (ESR) or C-reactive protein (CRP) or procalcitonin | 1 | 52 (100) | 70 (29.5) | <0.0001 |
| Sepsis, staphylococcal or streptococcal shock syndrome | -3 | 0(0) | 5(2.11) | - |

## Measures

Pearson’s Chi-square or Fisher’s exact test was applied wherever appropriate and a value of p <0.05 was considered as statistically significant. All the statistical analysis was performed using SPSS package version 19.0 (Armonk, NY: IBM Corp).

## Results

After screening and assessment of eligibility (Fig 1), a total of 1211 articles were identified through database searches and snowballing. Of these, 150 studies were found eligible for inclusion and analyzed in the systematic review. Of the 150 studies, 31 had aggregate data and hence were excluded. Finally, 119 studies with individual data were included.

**Fig 1:**
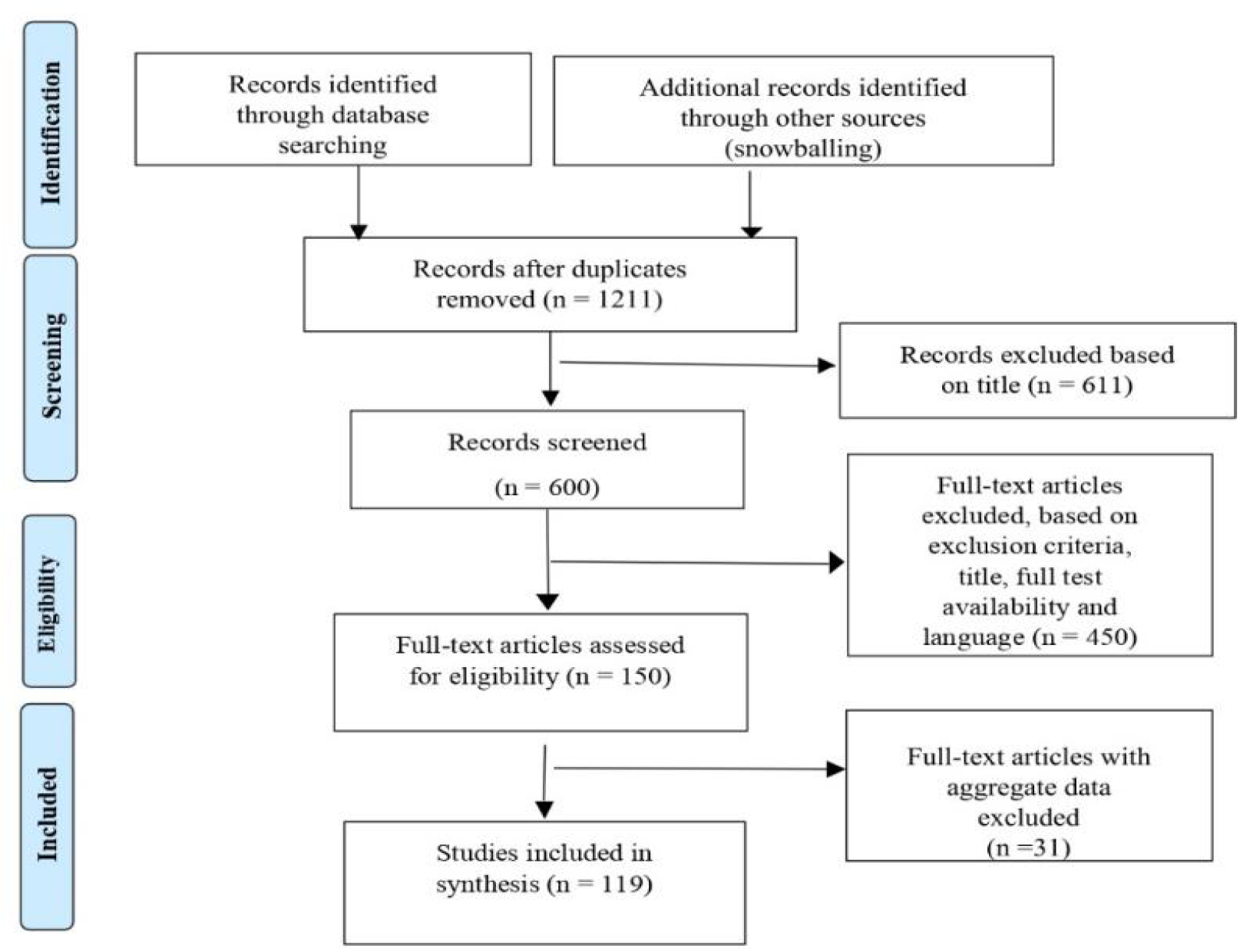
**PRISMA (Preferred Reporting Items for Systematic Review and Meta-analysis) flow chart for study selection**

### Study Characteristics

Data of 333 children reported from case series and case reports were compiled. Maximum studies were reported from China (n=42) followed by Italy (n=14), USA (n=16), Spain (n=6), France (n=6), UK (n=6) and Iran (n=5).

### Scoring and classification of MIS-C

Of the total number of children studied 44/333 (13.3%) were asymptomatic and 289/333 children (86.7%) were symptomatic. By applying scoring criteria (Table 1), 52/289 children (17.9%) could be classified to have MIS-C. All the 52 children were correctly classified as MIS-C by two authors blindly thereby validating the robustness of the scoring system. Interestingly, of the selected studies, only 25/52 cases were correctly classified as MIS-C as per WHO Criteria whereas 27 patients were identified as MIS-C as per the scoring system developed herein. Along with fever and raised markers of inflammation (Table 1), most of the children with MIS-C had cardiac involvement (80.8%), hypotension/shock (75%), GI manifestations (71.2%) and coagulopathy (55.8%) in significant proportion as compared to non MIS-C (p= <0.0001).

Existing comorbidities were reported in 17.3% (n=9/52) of children with MIS-C and 13.5% (n=32/237) without MIS-C. This difference was not statistically significant. In the MIS-C group, 63.5% of children (n = 33/52) required Intensive Care Unit (ICU) admission while only 15.2% (n = 36/237) children in the non-MIS-C group required ICU admission.This difference was statistically significant <0.0001.

As shown in Table 2, there were no sex biases in MIS-C, non-MIS-C and asymptomatic groups. Maximum numbers of SARS-CoV-2 positive children were reported in the age group of 5-14 years (Table 1) and this age group had a significantly higher proportion of children with MIS-C (p=0.004).

**Table 2:**
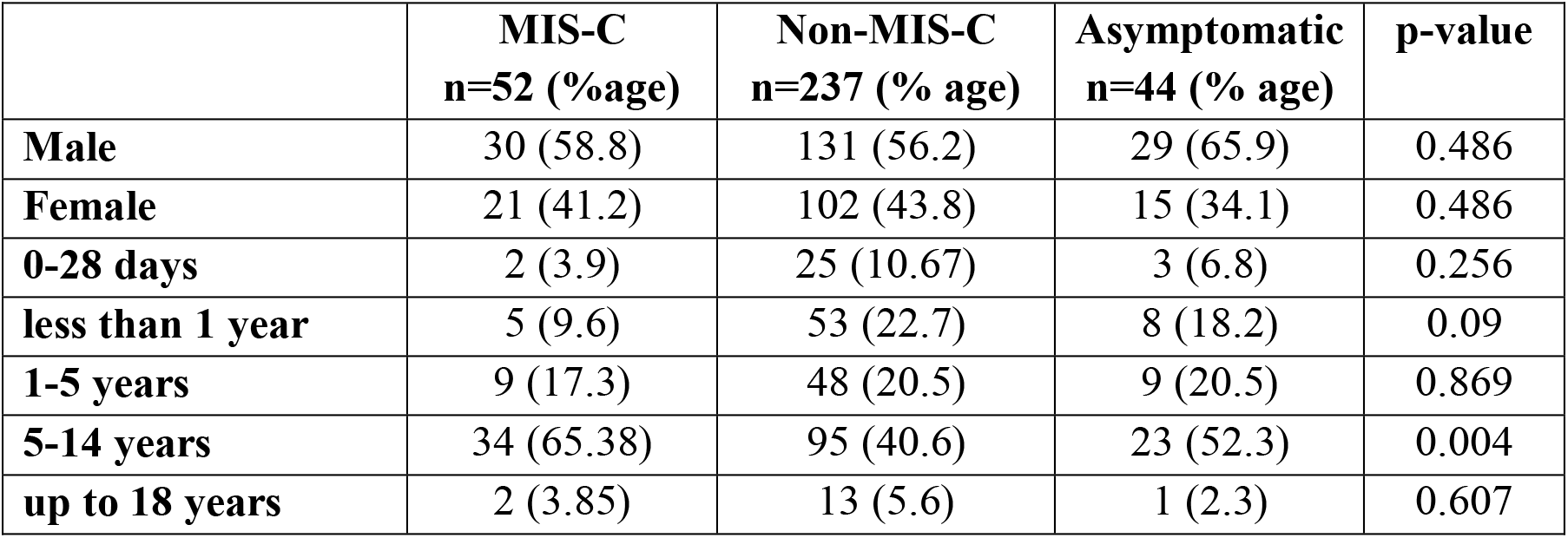
**Age and sex distribution of children with COVID-19 presenting with and without Pediatric Multisystem Inflammatory Syndrome (MIS-C) and asymptomatic presentation. The p-value of each variable (by Chi-square test) comparing MIS-C and Non-MIS-C groups**

### Presenting complaints

Table 3 compares the clinical presentation in children with and without MIS-C involvement. A significantly higher proportion of children with MIS-C reported fatigue/tiredness and hypotension as compared to children with non-MIS-C. Respiratory symptoms such as breathing difficulty and tachypnoea were significantly more in children with MIS-C. Cough and rhinorrhoea were less common in the MIS-C as compared to the non-MIS-C group.

**Table 3:** **Comparison of clinical presentation in children with or without Multisystem inflammatory Syndrome (MIS-C). The p-value is comparing MIS-C and Non-MIS-C**

MIS-C and COVID-19
| Presenting Complaints | MIS-C<br>n=52 (%) | Non-MIS-C<br>n=237 (%) | p-value |
| --- | --- | --- | --- |
| **General symptoms** |  |  |  |
| Fever | 52 (100) | 174 (73.4) | <0.0001 |
| Myalgia | 1 (1.9) | 4 (1.7) | 0.906 |
| Fatigue | 23 (44.2) | 20 (8.4) | <0.0001 |
| Loss of appetite/ reduced oral intake | 7 (13.5) | 6 (2.5) | 0.001 |
| Hypotension | 27 (51.9) | 5 (1.7) | <0.0001 |
| **Respiratory presentation** |  |  |  |
| Cough | 13 (25) | 119 (50.2) | 0.001 |
| Breathing difficulty | 24 (46.2) | 36 (15.2) | <0.0001 |
| Rhinorrhoea | 2 (3.8) | 54 (22.8) | 0.002 |
| Sore throat | 7 (13.5) | 27 (11.4) | 0.675 |
| Hypoxia | 29 (55.8) | 56 (23.6) | 0.029 |
| Apnoea | 1 (1.9) | 5 (2.1) | 0.932 |
| Tachypnoea | 25 (48.1) | 26 (10.6) | <0.0001 |
| Rhonchi/wheeze | 1 (1.9) | 5 (2.1) | 0.932 |
| Rales | 5 (9.6) | 11 (4.6) | 0.156 |
| **Cardiac presentation** |  |  |  |
| Cyanosis | 1 (1.9) | 7 (2.9) | 0.682 |
| Tachycardia | 35 (67.3) | 24 (9.7) | <0.0001 |
| Chest pain | 1 (1.9) | 4 (1.7) | 0.906 |
| Murmur | 1 (1.9) | 0 (0) | 0.032 |
| Abnormal ECG | 9 (17.3) | 0 (0) | <0.0001 |
| **Gastro-intestinal presentation** |  |  |  |
| Nausea/Vomiting | 29 (55.8) | 19 (8.0) | <0.0001 |
| Diarrhoea | 30 (57.7) | 22 (9.3) | <0.0001 |
| Dehydration | 24 (46.2) | 4 (1.7) | <0.0001 |
| Abdominal pain | 20 (38.5) | 4 (1.7) | <0.0001 |
| Intussusception | 0 (0) | 1 (0.4) | 0.639 |
| **Neurological presentation** |  |  |  |
| Headache | 8 (15.4) | 8 (3.38) | 0.001 |
| Drowsiness/irritability | 14 (26.9) | 21 (8.8) | <0.0001 |
| Convulsions | 5 (9.6) | 5 (2.1) | 0.007 |
| **Others** |  |  |  |
| Conjunctivitis | 11 (21.2) | 3 (1.3) | <0.0001 |
| Rash | 17 (32.7) | 7 (2.9) | <0.0001 |
| Skin lesions | 7 (13.5) | 2 (0.8) | <0.0001 |
| Lymphadenopathy | 3 (5.8) | 0 (0) | <0.0001 |
| Sepsis | 0 (0) | 6 (2.9) | 0.796 |
| Hematuria/Dysuria | 2 (3.9) | 2 (0.8) | 0.093 |

A significantly greater proportion of children with MIS-C had tachycardia and abnormal ECG on presentation. Vomiting, diarrhea, abdominal pain and dehydration were significantly more common in MIS-C group as compared to non-MIS-C group.

Headache and drowsiness/irritability were more prevalent in children with MIS-C as compared to non-MIS-C group. Convulsions were also reported in a greater proportion in children with MIS-C. Among children with MIS-C, 21% had conjunctivitis, 32.7% rash and 13.5% skin lesions, these were less common in children without MIS-C (3%, 7% and 2% respectively).

### Hematological changes

As shown in Table 4, a greater proportion of children in the MIS-C group had low hemoglobin as compared to the non-MIS-C group. Leukocytosis, neutrophilia and thrombocytopenia were more common in children with MIS-C as compared to children without MIS-C. However, neutropenia and lymphocytosis were only observed in the non-MIS-C group.

**Table 4:** **Comparision of Hematological investigations and Cardiac dysfunction in children with and without Pediatric Multisystem inflammatory Syndrome (MIS-C)**

|  | MIS-C (n=52) |  | Non-MIS-C (n=237) |  |  |
| --- | --- | --- | --- | --- | --- |
| Laboratory investigations | Tests results available | n (%) | Tests results available | n (%age) | p-value |
| **Complete blood counts** |  |  |  |  |  |
| ↓ Hb | 48 | 10 (20.8) | 195 | 11 (5.6) | 0.01 |
| ↑ WBC | 48 | 13 (27.1) | 194 | 21 (10.8) | 0.004 |
| ↓ WBC | 48 | 13 (27.1) | 194 | 38 (19.6) | 0.460 |
| ↑ ANC | 48 | 20 (41.7) | 192 | 11 (5.7) | <0.0001 |
| ↓ ANC | 48 | 0 (0.0) | 192 | 43 (22.4) | <0.0001 |
| ↑ ALC | 48 | 0 (0.0) | 192 | 3 (18.8) | 0.001 |
| ↓ ALC | 48 | 26 (54.2) | 192 | 57 (29.7) | 0.001 |
| ↓ Platelets | 48 | 16 (33.3) | 195 | 11 (5.6) | <0.0001 |
| **Inflammatory Markers** |  |  |  |  |  |
| ↑ ESR | 30 | 15 (46.9) | 127 | 11 (8.7) | <0.0001 |
| ↑ CRP | 52 | 49 (96.1) | 152 | 44 (29.0) | <0.0001 |
| ↑ Procalcitonin | 29 | 25 (86.2) | 78 | 39 (50.0) | 0.001 |
| ↑ Ferritin | 31 | 24 (77.4) | 13 | 4 (30.8) | 0.003 |
| ↑ LDH | 12 | 8 (66.7) | 74 | 27 (36.5) | 0.048 |
| ↑ Fibrinogen | 13 | 10 (77.0) | 18 | 1 (5.6) | <0.0001 |
| ↑ IL-6 | 17 | 14 (82.4) | 23 | 12 (52.2) | 0.048 |
| **Coagulation markers** |  |  |  |  |  |
| ↑ APTT | 5 | 3 (60) | 31 | 1 (3.2) | <0.0001 |
| ↑ PT | 6 | 4 (66) | 33 | 0 (0.0) | <0.0001 |
| ↑ D-Dimer | 23 | 22 (95.6) | 39 | 10 (34.5) | <0.0001 |
| **Other investigations** |  |  |  |  |  |
| ↑ AST/SGOT | 32 | 14 (43.8) | 75 | 13 (17.3) | 0.004 |
| ↑ ALT/SGPT | 32 | 14 (43.8) | 81 | 15 (18.5) | 0.006 |
| ↑ Urea | 26 | 7 (27.0) | 63 | 0 (0.0) | <0.0001 |
| ↑ Creatinine | 26 | 10 (38.5) | 64 | 5 (4.7) | <0.0001 |
| ↑ CK | 35 | 5 (29.4) | 179 | 10 (17.2) | 0.270 |
| ↑ CK-MB | 49 | 1 (33.3) | 211 | 1 (3.9) | 0.056 |
| ↑ Troponin | 14 | 31 (81.6) | 223 | 1 (7.1) | <0.0001 |
| ↑ Myoglobin | 50 | 0 (0.0) | 234 | 1 (33.3) | 0.361 |
| ↑ Pro-BNP | 28 | 24 (100) | 234 | 0 (0.0) | <0.0001 |
| Abnormal echocardiogram | 10 | 26 (70.0) | 229 | 4 (66.7) | 0.858 |

**Abbreviations:** **Hb:** Hemoglobin **WBC:** Whole blood counts **ANC:** Absolute Neutrophils count **ALC:** Absolute Lymphocyte Count **CRP:** C Reactive Protein **LDH:** Lactic acid Dehydrogenase **IL-6:** Interleukin 6. **APTT:** Activated Partial Thromboplastin time **PTT:** Partial Thromboplastin Time **AST/SGOT:** Aspartate aminotransferase/serum glutamic oxaloacetic transaminase **ALT/SGPT:** alanine aminotransferase/ serum glutamic-pyruvic transaminase **CK:** Creatinine Kinase **CK-MB:** Creatine kinase myocardial band **Pro-BNP:** N-terminal pro-Brain Natriuretic Peptide **ABN ECHO:** Abnormal Echocardiography

Amongst the inflammatory markers (Table 4), elevated Erythrocyte Sedimentation Rate (ESR), serum C-Reactive Protein (CRP) and Procalcitonin were found in significantly higher proportion of children with MIS-C as compared to non-MIS-C. Similarly, a significantly greater proportion of children with MIS-C had elevated serum Ferritin, Lactate Dehydrogenase (LDH), Fibrinogen and IL-6 as compared to their non-MIS-C counterparts.

Elevated levels of coagulation markers like prothrombin time (PT), partial thromboplastin time PTT) and D-dimer were found in significantly greater proportion of children with MIS-C as compared to non-MIS-C group (Table 4). A significantly greater proportion of children in the MIS-C group had raised levels of liver function markers compared to the non-MIS-C group. Elevated levels of renal function markers were observed in a significantly higher proportion of children with MIS-C as compared to non-MIS-C groups.

### Cardiac dysfunction

Amongst cardiac enzymes, creatinine kinase-MB (CK-MB), Troponin and N-terminal pro-Brain Natriuretic Peptide were elevated in a significantly higher proportion of children with MIS-C. Echocardiography was abnormal in 26 children with MIS-C and 4 children without MIS-C.

### Radiographic changes

Chest X-ray abnormalities were reported in 42.3% of children with MIS-C and 24% of children without MIS-C (p=0.007). As compared to MIS-C group, a significantly greater proportion of children without MIS-C had abnormal HRCT (High-Resolution Computer Tomography) and this difference was statistically significant (p=0.0001).

### Clinical Management

Antiviral medications such as Oseltamivir, Lopinavir/Ritonavir, and Ribavirin were used in a higher proportion in the non-MIS-C group as compared to the MIS-C group. Methylprednisolone was more commonly used in children with MIS-C as compared to those without MIS-C. Immunoglobulins were used in 63.5% of children with MIS-C and 3.4% in children without MIS-C. Tocilizumab was used in 4 children with MIS-C and 2 children without MIS-C. Use of Infliximab was reported in 2 children with MIS-C (Table 5).

**Table 5:** **Comparison of management strategies in children with COVID-19 with or without Multisystem inflammatory Syndrome in Children (MIS-C)**.

|  | MIS-C n=52 (%) | Non-MIS-C n=237 (%) | P-value |
| --- | --- | --- | --- |
| **Antiviral Medications** | 9 (17.3) | 68 (28.7) | 0.093 |
| **Steroids** | 27 (52.0) | 11 (4.7) | < 0.00001 |
| **IV Immune globulin** | 33 (63.5) | 8 (3.4) | < 0.00001 |
| **Interferon -1B Inhalation** | 3 (5.8) | 37 (15.6) | 0.061 |
| **Tocilizumab and Infliximab** | 5 (9.7) | 2 (0.9) | 0.003 |
| **Anticoagulation** |  |  |  |
| **Aspirin** | 14 (26.9) | 0 (0.0) | < 0.00001 |
| **IV anticoagulation** | 11 (21.1) | 1 (0.4) | < 0.00001 |
| **Respiratory support** |  |  |  |
| **Oxygen** | 39 (75) | 49 (20.7) | < 0.00001 |
| **Non invasive ventilation** | 11 (21.2) | 9 (3.8) | < 0.00001 |
| **Mechanical Ventilation** | 17 (32.7) | 6 (2.5) | < 0.00001 |
| **High flow ventilation** | 0 (0) | 1 (0.4) | 1.000 |
| **Inotropes** | 28 (53.8) | 2 (0.8) | < 0.00001 |
| **Hydroxychloroquine** | 11 (21.2) | 17 (7.2) | 0.002 |
| **Antibiotic** | 40 (76.9) | 67 (28.9) | < 0.00001 |
| **Miscellaneous** |  |  |  |
| **ECMO, Plasmapheresis** | 3 (5.8) | 0 (0.0) | 0.0056. |
| **Discharged** | 32 (61.0) | 219 (92.4) | < 0.00001 |
| **Mortality** | 5 (9.6) | 1 (0.4) | 0.001 |

*ECMO- Extra corporeal membrane oxygenation*

The use of intensive modalities was significantly higher in children with MIS-C. Oxygen supplementation, non-invasive ventilation, mechanical ventilation and inotropes were commonly used in children with MIS-C.

Aspirin and intravenous anticoagulation usage were reportedly used in children with MIS-C; Hydroxychloroquine was used in 21.2% of children with MIS-C and 7.2 % of children without MIS-C. Antibiotic use was reported in 77% of children with MIS-C and 67% of children without MIS-C (Table 5). Mortality rates were significantly higher in children with MIS-C (Table 5).

## Discussion

The present systematic review compared the clinical presentation, laboratory parameters and management strategies used in symptomatic SARS-CoV-2 positive children with and without MIS-C. The results reveal that the COVID-19 children with MIS-C have a more severe presentation, higher adverse outcomes and required aggressive management modalities as compared to their non-MIS-C counterparts.

Several studies and systematic reviews have been published describing the clinical presentations and outcomes of children with COVID-19 presenting with MIS-C^18–21^. However, a major limitation of the systematic reviews is that the clinical case definition of MIS-C was as per the author criteria which vary significantly between different studies. Further, as definition of MIS-C is an evolving concept, many studies have misclassified the cases in absence of an objective criteria. To attain objectivity and uniformity across studies we first evolved a scoring system for the clinical signs of MIS-C as per WHO criteria. We gave the highest weightage to fever as it is a mandatory requirement to classify MIS-C as per WHO criteria. As not all children will present with all the symptoms, a score of 1 was given for other parameters. As septicemia can also present like MIS-C^22^, it is necessary to rule out sepsis in the children. A high negative score for sepsis was thus included to provide an accurate differential diagnosis of COVID-19 associated MIS-C. This scoring system was implemented on the primary data collected for 333 children and two authors accurately classified 52 children having MIS-C. Although this scoring needs to be clinically validated in a hospital setup, we believe that our scoring system is robust and can be utilized by clinicians to classify MIS-C.

Overall our study estimated the prevalence of MIS-C in 21% of children with symptomatic COVID-19. We next used our scoring system to compare the presentation of 52 children with MIS-C vs 237 non-MIS-C cases. We noted that the risk of MIS-C is low (∼10%) in children with COVID-19 below one year of age which increased to almost 17% in the age group of 1-5 years. However, a large number of children (65%) with COVID-19 in the age group of 5-14 years developed MIS-C. Similar to observational studies^12,23^, our systematic review shows that the disease is of higher preponderance in the age group of 5-14 years. This age distribution is different from that of classical Kawasaki disease which is common in children of younger age^24^. Thus, children in the age group of 5-14 years developing symptomatic COVID-19 disease must be closely monitored for progression towards MIS-C.

It was observed that beyond inflammation and fever, the children with MIS-C had more severity at presentation. Almost 75-80% of the children with MIS-C and COVID-19 had gastrointestinal involvements which were rarely seen in COVID-19 children without MIS-C involvement. Clinicians should be alert to notice signs and symptoms such as nausea/vomiting, abdominal pain and diarrhea for an impending MIS-C.

We observed that cardiac dysfunctions are very common in the MIS-C group as compared to non-MIS-C group. Cardiac dysfunctions in the form of tall and wide P waves, ST-segment abnormalities, T negative waves are reported in a proportion of children with COVID-19^14,24–26^. However, coronary artery involvement in form of diffuse ectasia and dilatation was reported in relatively fewer cases of children with COVID-19^11,28–30^. It is known that cardiac enzymes such as CK, CK-MB, Troponin and N-terminal pro-Brain Natriuretic Peptide are elevated in a proportion of children with COVID-19^11,12,26,30,31^. We observed that these markers were mostly elevated in children with MIS-C and not so in the non-MIS-C group. Neurological defects are reported in some cases of children with COVID-19^26,32–37^. Herein we observed a reasonable preponderance of neurological involvement such as headache, convulsions encephalitis and meningoencephalitis in the MIS-C group implying the need for follow-up for long-term neurological deficits in children with COVID-19. Thus, while managing pediatric cases of COVID-19, clinicians must pay attention to symptoms such as encephalitis, headache or convulsions for possible development of MIS-C.

Serum biochemistry is indispensable in the differential diagnosis of many conditions in children. Many biochemical parameters such as inflammatory markers, cardiac and liver enzymes are reportedly altered in children with MIS-C. More than 80% of children with MIS-C had raised IL-6 which is an important marker for cytokine storm^14,25,29,33,38–40^. Our study revealed that the traditional inflammatory markers such as raised ESR, CRP, IL-6 and Procalcitonin are observed in children with MIS-C. However, for these markers to be diagnostic of MIS-C, septicemia should be ruled out which often is time-consuming and reports may not be readily available. Based on our study we propose that elevated coagulation markers (PTT, PT, and D-Dimers) along with elevated cardiac markers such as Troponin and pro-BNP could be useful surrogates to identify MIS-C in COVID-19 children with MIS-C. Larger clinical studies will be required to determine the sensitivity and specificity of these markers for their diagnostic use in COVID-19 related MIS-C.

Children with COVID-19 and MIS-C required aggressive management strategies. Intensive care admissions were significantly more common in MIS-C group as compared with children without MIS-C presentation. Based on the presenting phenotypes, authors reported the use of antibiotics, steroids, immunoglobulins and inotropes in children with MIS-C. Even though septicemia is an exclusion criterion for MIS-C, antibiotics use was reported in 77% of cases with COVID-19 related MIS-C. Clinicians must be aware of the fact that bacterial septicemia is rare in children with COVID-19 and MIS-C and hence antibiotics will be ineffective and should be avoided. Use of Tocilizumab and Infliximab was reported in children with MIS-C with elevated IL-6 levels^14,38,40^. Use of recombinant IL-1 receptor antagonist (Anakinra) was reported in a study from UK with high IL-1 values^29^. Anticoagulation therapy such as prophylactic low molecular weight Heparin and Enoxaparin was mainly reported in case of severe cardiac involvement^14,25,41^. Aspirin use was mainly reported in children with Kawasaki like presentation^12,42–44^. These observations point towards evolving need for prophylactic anticoagulation in the treatment of MIS-C. Unfortunately, in absence of the data on the effectiveness of these therapies, it is difficult to comment which of these could be beneficial. The recovery rate was higher in the non-MIS-C group and overall mortality was only 1.8%, while the mortality was almost 10% in children with MIS-C. Although the numbers of children are fewer, this preliminary analysis suggests that MIS-C is associated with high morbidity and mortality

Although, the presentations of MIS-C in children with COVID-19 strongly overlap with that of Kawasaki disease, clinically MIS-C seems to significantly differ from classical KD. MIS-C commonly affects older children and adolescents, whereas classic KD typically affects infants and young children. The diagnosis of KD requires the presence of fever lasting at least 5 days with at least 4 of the 5 signs of mucocutaneous inflammation. Amongst these, oral mucous membrane involvement, polymorphous rash, extremity changes, ocular changes, and cervical lymphadenopathy are common in KD^45–48^. Other findings such as gastrointestinal symptoms are common in MIS-C compared to classic KD^24,49^. Additionally, approximately 30% of patients with KD have dilatation of coronary arteries^50,51^. Amongst the hematological parameters, decreased hemoglobin, lymphopenia thrombocytopenia along with elevated inflammatory markers were characteristically reported in MIS-C contrary to thrombocytosis with lymphocytosis which is more evident in typical KD^12,14,43,44,52–54,26,28– 31,39,40,42^.

Similarly, differentiating MIS-C and acute COVID-19 is also crucial. The patterns of clinical presentation and organ system involvement differentiate MIS-C from severe acute COVID-19^55,56,57^. Severe pulmonary involvement (pneumonia and acute respiratory distress syndrome) is characteristic of severe acute COVID-19 as it is more common in cases without MIS-C. In cases with MIS-C, it is often secondary to cardiac dysfunction. Further, myocardial dysfunction, muco-cutaneous and gastrointestinal involvement are more common in MIS-C than in severe acute COVID-19. Elevated inflammatory markers, lymphopenia and thrombocytopenia tend to be more common in MIS-C. SARS-CoV-2 antibody titres may also be higher in patients with MIS-C compared with acute COVID-19^58^. Although all these need to be systematically investigated to determine the accurate clinical and laboratory markers for a differential diagnosis of MIS-C and acute COVID-19 in children.

## Conclusion

Multisystem inflammatory involvement in pediatric COVID-19 is a distinct entity and is not rare as previously thought. The clinical spectrum of MIS-C appears to differ from typical Kawasaki presentation reiterating that MIS-C could be a peculiar phenomenon of COVID-19. In the background of increasing diversity of COVID-19 presentation in pediatric age group and in the wake of second wave of COVID-19, timely diagnosis and management of MIS-C is utmost importance. The scoring proposed in this review may be adapted for meticulous identification of children with MIS-C in SARS-CoV-2 infection. The information generated in this review should act as a primer to consider MIS-C as a distinct presentation of COVID-19 in pediatric age groups. The results will aid in decision-making for devising diagnostics and treatment modalities for MIS-C and Covid-19.

## Data Availability

The data is available with all the authors.

## Ethical Approval

The present systematic review is under exempt category as per Institutional Ethics Committee (ID-ICEC/Sci-50/53/2020).

## Acknowledgments

The study is supported by grants from the Indian Council of Medical Research (ICMR) intramural funding to NIRRH. The manuscript bears ICMR-NIRRH ID RA/948/07-2020.

## Author contribution

SS, SM, and DM conceived the study. SS and SJ collected and analyzed the literature and shortlisted the studies. SS extracted the data and RG verified it. SM coordinated the study.

